# Emergence of Clade III *Candida auris* of African Origin in Los Angeles

**DOI:** 10.1101/2020.10.26.20214908

**Authors:** Travis K. Price, Ruel Mirasol, Kevin W. Ward, Ayrton J. Dayo, Sukantha Chandrasekaran, Omai B. Garner, Annabelle de St Maurice, Shangxin Yang

**Author notes:** **CORRESPONDING AUTHOR:** Shangxin Yang, PhD, D(ABMM); UCLA Clinical Microbiology Laboratory, 11633 San Vicente Blvd, Los Angeles, CA 90049.

## Abstract

*Candida auris* is an emerging multi-drug resistant yeast responsible for invasive infections among hospitalized patients. We described a case series of *C. auris* isolated in the Los Angeles area from an ongoing outbreak. Genomic analysis showed a single Clade III lineage of African origin in all 6 cases since 2019.

## Introduction

*Candida auris* was first isolated in 2009 from the external ear canal of a 70-year-old patient in Tokyo, Japan^1^. In the decade that followed, cases of *C. auris* bloodstream and other invasive infections have been reported worldwide^2 3 4 5 6 7^. Many strains of *C. auris* are multidrug resistant and in some cases have elevated minimum inhibitory concentrations (MICs) to all three classes of antifungals including azoles, echinocandins and polyenes. The Centers for Disease Control and Prevention (CDC) recently listed *C. auris* as one of five “urgent threats” to public health in the 2019 Antibiotic Resistance (AR) Threats Report^8^, highlighting the need for active surveillance and appropriate infection prevention.

Whole genome sequencing (WGS) and phylogenetic analyses revealed the presence of at least four major clades of *C. auris* with worldwide geographic associations^9 10^. In the United States, outbreaks have been documented in New York, New Jersey, and Illinois; these were generally caused by strains of *C. auris* that were part of Clades I and IV^11^.

Here, we describe several cases of *C. auris* colonization and infection in patients of long-term acute care (LTAC) facilities in the Los Angeles area. WGS revealed that all isolates were part of Clade III, likely representing a unique outbreak in the United States. This is the first description of *C. auris* strains in Southern California.

## Methods

A laboratory-developed polymerase chain reaction (PCR) test implemented by the clinical microbiology laboratory in September 2019, was used for the detection of *C. auris* from inguinal-axillary swabs and yeast isolates from positive fungal culture of clinical specimens as a means to actively screen high-risk patients transferred to UCLA from LTAC facilities and skilled nursing facilities (SNFs). Nucleic acid extraction using the EZ1 Tissue Extraction kit (Qiagen, Germantown, MD) was performed followed by PCR using the DiaSorin Molecular LIAISON^®^ MDX platform (DiaSorin Molecular, Cypress, CA). PCR-positive swab specimens were cultured; yeast isolates were identified using VITEK MS MALDI-TOF system (bioMérieux Inc, Hazelwood, MO).

Antifungal susceptibility testing by broth microdilution was performed on panels prepared in-house according to Clinical and Laboratory Standards Institute (CLSI) standards. The CDC tentative MIC breakpoints were used for interpretation of the following drugs: amphotericin B (≥2µg/ml), fluconazole (≥32µg/ml), anidulafungin (≥4µg/ml), caspofungin (≥2µg/ml), micafungin (≥4µg/ml). Voriconazole, itraconazole, and posaconazole were also tested but no interpretative criteria for *C. auris* are available.

WGS was performed using Illumina MiSeq (Illumina, San Diego, CA). Analyses were performed using CLC Genomics Workbench (Qiagen, Valencia, CA, USA) and Geneious Prime (Biomatters, New Zealand). Reads were mapped to a reference *C. auris* complete genome (ATCC MYA-5002, NCBI: CP043531); this strain is part of Clade III. Sequences were compared by K-mer analysis and single nucleotide polymorphism (SNP) analysis using maximum-likelihood phylogenies.

This is a retrospective analysis and is part of an infection prevention and quality improvement project. The institutional review boards (IRB) approval was waived by the UCLA Office of the Human Research Protection Program.

## Results

In one year (09/2019 – 09/2020), we screened 113 patients using our in-house *C. auris* PCR assay; six patients have tested positive for *C. auris*. Cycle threshold (Ct) values ranged from 22.6 to 39.7 (**Table 1**). Of the six PCR-positive patients, herein referred to as patients A through F, five tested positive between July and September 2020 (patients B-F); while patient A tested positive in October 2019.

**Table 1.** Patient test results, clinical history, and *C. auris* isolate information.

| Patient | Month of Positive PCR | PCR Ct Value | *C. auris* Isolate (Specimen Type) |
| --- | --- | --- | --- |
| A | 10/2019 | 22.6 | UCLA_A1 (Inguinal-Axillary)<br>UCLA_A2 (Tracheal) |
| B | 7/2020 | 39.5 | Not isolated<sup>a</sup> |
| C | 8/2020 | 22.6 | UCLA_C1 (Inguinal-Axillary) |
| D | 8/2020 | 39.7 | UCLA_D1 (Inguinal-Axillary) |
| E | 8/2020 | 28.3 | UCLA_E1 (Inguinal-Axillary) |
| F | 9/2020 | 30.6 | UCLA_F1 (Pleural Fluid, Rt) |

<sup>a</sup> *C. auris* was not isolated from the inguinal-axillary surveillance swab of patient B

All six patients were residents of 4 different LTAC facilities within the Los Angeles county area; all had history of tracheostomy. Patients A and F had prior history of *C. auris* colonization. The microbiological findings of each patient are described in **Table 1**. *C. auris* isolates grew from inguinal-axillary swabs of 4/6 patients (A, C, D, E) and from other clinical specimens of patients A (tracheal aspirate) and F (pleural fluid). We did not isolate *C. auris* from patient B (Ct=39.5).

All *C. auris* isolates were resistant to amphotericin B (MIC=2µg/ml) and fluconazole (MIC>64µg/ml) but susceptible to echinocandins (**Table 2**).

**Table 2.** Antifungal susceptibility results for *C. auris* isolates.

| Antifungal | MIC (µg/ml) (interpretation) for *C. auris* isolate: |  |  |  |  |  |
| --- | --- | --- | --- | --- | --- | --- |
|  | UCLA_A1 | UCLA_A2 | UCLA_C1 | UCLA_D1 | UCLA_E1 | UCLA_F1 |
| Amphotericin B | 2 (R) | 2 (R) | 2 (R) | 2 (R) | 2 (R) | 2 (R) |
| Fluconazole | >64 (R) | >64 (R) | >64 (R) | >64 (R) | >64 (R) | >64 (R) |
| Voriconazole | 2 | 1 | 1 | 0.5 | 0.5 | 2 |
| Itraconazole | 1 | 1 | 0.5 | 0.25 | 0.25 | 1 |
| Posaconazole | 0.06 | 0.12 | ≤0.03 | ≤0.03 | 0.06 | 0.06 |
| Anidulafungin | 0.5 (S) | 0.12 (S) | 0.25 (S) | 0.12 (S) | 0.06 (S) | 1 (S) |
| Caspofungin | 0.5 (S) | 0.12 (S) | 0.5 (S) | 0.5 (S) | 0.12 (S) | 0.5 (S) |
| Micafungin | 0.25 (S) | 0.25 (S) | 0.25 (S) | 0.12 (S) | 0.25 (S) | 0.25 (S) |

MIC testing was performed on panels prepared in-house in accordance with CLSI guidelines. Interpretive breakpoints were defined by the CDC Antifungal Susceptibility Testing and Interpretation guidelines for *C. auris*, which were adapted from interpretive criteria for closely related *Candida spp.* Tentative breakpoints for the following antifungals were used: amphotericin B (≥2µg/ml), fluconazole (≥32µg/ml), anidulafungin (≥4µg/ml), caspofungin (≥2µg/ml), micafungin (≥4µg/ml). S=susceptible, I=intermediate, R=resistant.

K-mer analysis was performed using 252 publicly available sequences of *C. auris* described previously^12^ (**Supplemental Data 1**) in addition to the sequences of the six isolates described here. All UCLA isolates belonged to Clade III (**Supplemental Data 1**). In this analysis, Clade III contained 56 isolates; these were primarily isolated from the African continent (n=44)^11^ (**Supplemental Figure 1**).

Isolates from all four clades have been identified in the US and show a geographic relationship^11^; Clade I is predominant. A k-mer phylogenetic analysis of 57 *C. auris* isolated in the US was performed (**Figure 1**). Clade III (n=7), which included the six UCLA isolates, had one additional isolate which originated from Indiana and was isolated in March of 2017 (Genbank accession # SRR7909359). SNP analysis showed that the UCLA isolates were closely related to one another (3-78 SNPs) as well as to the Indiana isolate (66 – 138 SNPs) (**Supplemental Table 1**).

**Figure 1.**
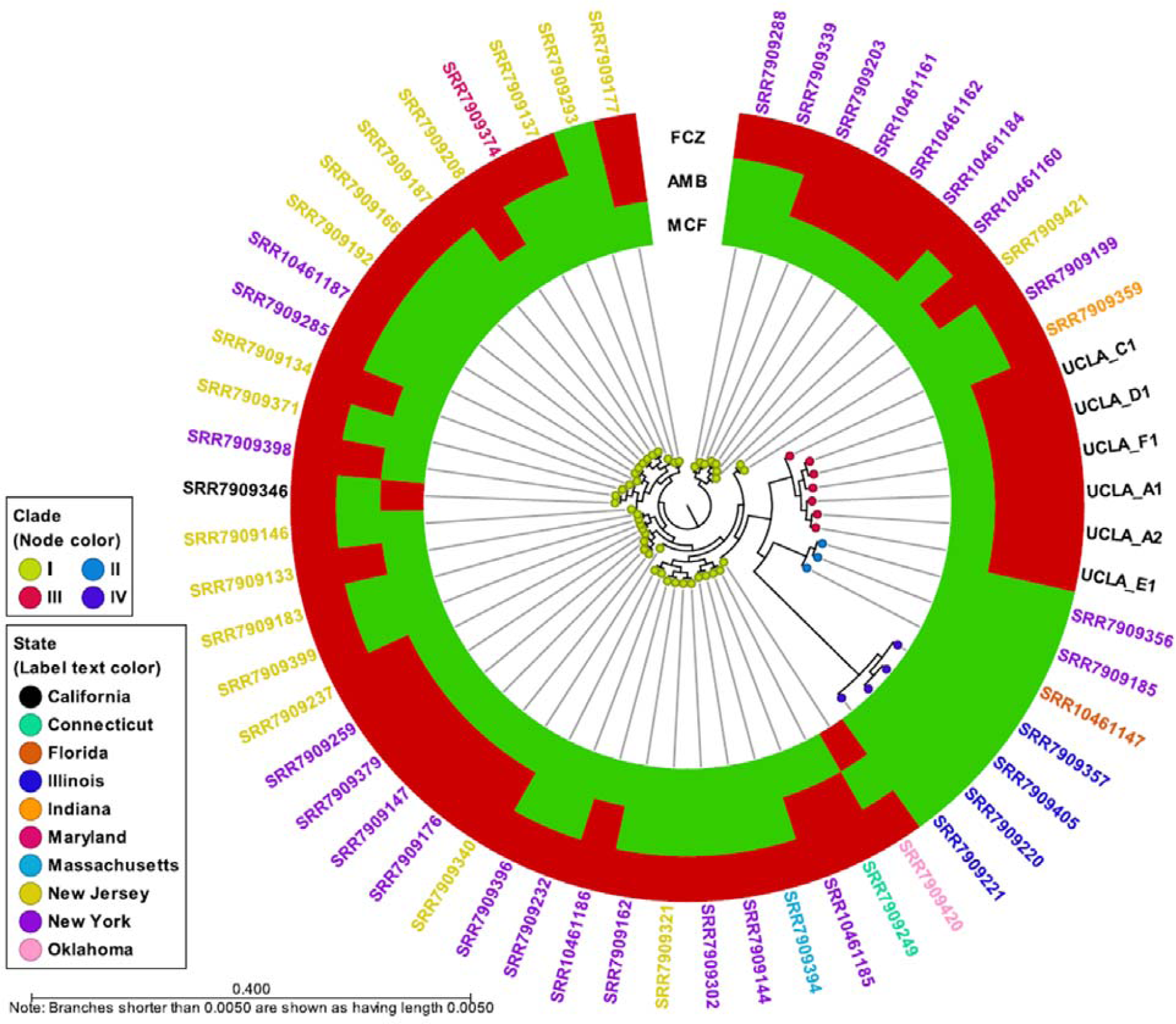
K-mer Analysis of 57 *C. auris* Isolates from the United States. K-mer analysis was performed using CLC Genomics Workbench (Qiagen, Valencia, CA, USA) using genome sequences from 52 publicly available *C. auris* strains on NCBI GenBank as well as sequences from UCLA_A1, UCLA_A2, UCLA_C1, UCLA_D1, UCLA_E1, and UCLA_F1. A list of sequences used is found in Supplemental Data 1. Each node represents a unique isolate; the node color refers to the clade. The color of the isolate name (*i*.*e*., label text color) refers to the state of origin. The metadata shows the susceptibility of each isolate to fluconazole (FCZ), amphotericin B (AMB), and micafungin (MCF); red indicates resistant, green indicates susceptible.

To further characterize the *C. auris* isolates, we analyzed the sequence of two genes associated with antifungal resistance: *erg11*, which encodes lanosterol 14-alpha demethylase, and *fks1*, a subunit of 1,3-β-D-glucan synthase. Sequences of *erg11* were identical for all UCLA isolates; all had a 99.6% pairwise nucleotide identity to the reference (CP043531) and two amino acid substitutions: V125A and F126L (**Supplemental Table 2**). Mutations at amino acid 126 are associated with increased azole resistance in *C. auris* ^9^ and are a common feature of Clade III isolates^13^. The specific F126L mutation appears to be exclusive to Clade III^12^. These findings are consistent with the antifungal susceptibility testing which showed that all isolates were resistant to fluconazole (**Table 2**). Sequences of *fks1* were identical in five of the UCLA isolates (A1, A2, C1, D1, E1) and showed 99.9% pairwise nucleotide identity to the reference (CP043531); these isolates had one amino acid substitution: I1572L (**Supplemental Table 2**). Isolate UCLA_F1 had the same substitution in addition to I1095L. All isolates had a WT serine at amino acid 639; mutations at position 639 are linked to echinocandin resistance in *C. auris*^14^. All isolates were susceptible to caspofungin, micafungin and anidulafungin.

## Discussion

Here, our use of an in-house PCR test to actively screen patients for *C. auris* colonization has proven to be vital in identifying and preventing the spread of this pathogen in our hospital system. WGS of isolates obtained from several patients of LTAC facilities revealed two important findings. First, these isolates are closely related to one another; this confirms the existence of an ongoing outbreak with community spread in the Los Angeles area. Second, these isolates are part of Clade III of African origin; this suggests a unique introduction of *C. auris* into the United States because this clade has not been implicated in outbreaks in other states.

The isolates described here were all resistant to fluconazole and Amphotericin B but susceptible to echinocandins. Additionally, all isolates had an F126L mutation in the *erg11* gene, which is associated with fluconazole resistance and unique to Clade III strains of *C. auris*.

Limitations of this study include the lack of any additional epidemiological history of the patients related to exposures including travel. Being able to link patients to a country with known *C. auris* outbreaks of Clade III strains would be essential to understanding how the current Los Angeles outbreak began.

In conclusion, we have identified a unique Clade III *C. auris* strain in an ongoing outbreak in the LTAC facilities since 2019, indicating active community spread of multi-drug resistant *C. auris* in the Los Angeles area.

## Data Availability

We are in the process of uploading the WGS data of the C. auris to NCBI.

## SUPPLEMENTAL FIGURES & TABLES

**Supplemental Table 1.** SNP analysis of *C. auris* isolates.

|  | UCLA_A1 | UCLA_A2 | UCLA_C1 | UCLA_D1 | UCLA_E1 | UCLA_F1 | US Clade III<br>(SRR7909359) | US Clade I<br>(SRR7909162) | US Clade II<br>(SRR7909356) | US Clade IV<br>(SRR7909221) |
| --- | --- | --- | --- | --- | --- | --- | --- | --- | --- | --- |
| UCLA_A1 | --- | 3 | 12 | 10 | 7 | 76 | 66 | 24373 | 35804 | 95894 |
| UCLA_A2 | 3 | --- | 11 | 9 | 6 | 77 | 65 | 24372 | 35803 | 95893 |
| UCLA_C1 | 12 | 11 | --- | 4 | 7 | 76 | 74 | 24381 | 35808 | 95896 |
| UCLA_D1 | 10 | 9 | 4 | --- | 5 | 78 | 72 | 24379 | 35810 | 95897 |
| UCLA_E1 | 7 | 6 | 7 | 5 | --- | 75 | 69 | 24376 | 35807 | 95897 |
| UCLA_F1 | 76 | 77 | 76 | 78 | 75 | --- | 138 | 24402 | 35796 | 95919 |
| US Clade III<br>(SRR7909359) | 66 | 65 | 74 | 72 | 69 | 138 | --- | 24384 | 35814 | 95909 |
| US Clade I<br>(SRR7909162) | 24373 | 24372 | 24381 | 24379 | 24376 | 24402 | 24384 | --- | 36609 | 97646 |
| US Clade II<br>(SRR7909356) | 35804 | 35803 | 35808 | 35810 | 35807 | 35796 | 35814 | 36609 | --- | 95704 |
| US Clade IV<br>(SRR7909221) | 95894 | 95893 | 95896 | 95897 | 95897 | 95919 | 95909 | 95704 | 97646 | --- |

Reads were mapped to the *C. auris* reference genome (CP043531), consensus sequences were extracted from the alignments and SNP analysis was performed.

**Supplemental Table 2.** Nucleotide and amino acid variants of *C. auris* isolates.

| *C. auris*<br>Isolate | Erg11 |  | Fks1 |  |
| --- | --- | --- | --- | --- |
|  | Nucleotide<br>Pairwise Identity | Protein Sequence<br>Variants | Nucleotide<br>Pairwise Identity | Protein Sequence<br>Variants |
| UCLA_A1 | 99.6% | p.V125A<br>p.F126L | 99.9% | p.I1572L |
| UCLA_A2 | 99.6% | p.V125A<br>p.F126L | 99.9% | p.I1572L |
| UCLA_C1 | 99.6% | p.V125A<br>p.F126L | 99.9% | p.I1572L |
| UCLA_D1 | 99.6% | p.V125A<br>p.F126L | 99.9% | p.I1572L |
| UCLA_E1 | 99.6% | p.V125A<br>p.F126L | 99.9% | p.I1572L |
| UCLA_F1 | 99.6% | p.V125A<br>p.F126L | 99.8% | p.I1095L<br>p.I1572L |

Nucleotide pairwise identity to *C. auris* strain ATCC MYA-5002 (CP043531), and protein translation and variant analysis was performed using Geneious Prime (Biomatters, New Zealand).

**Supplemental Figure 1.**
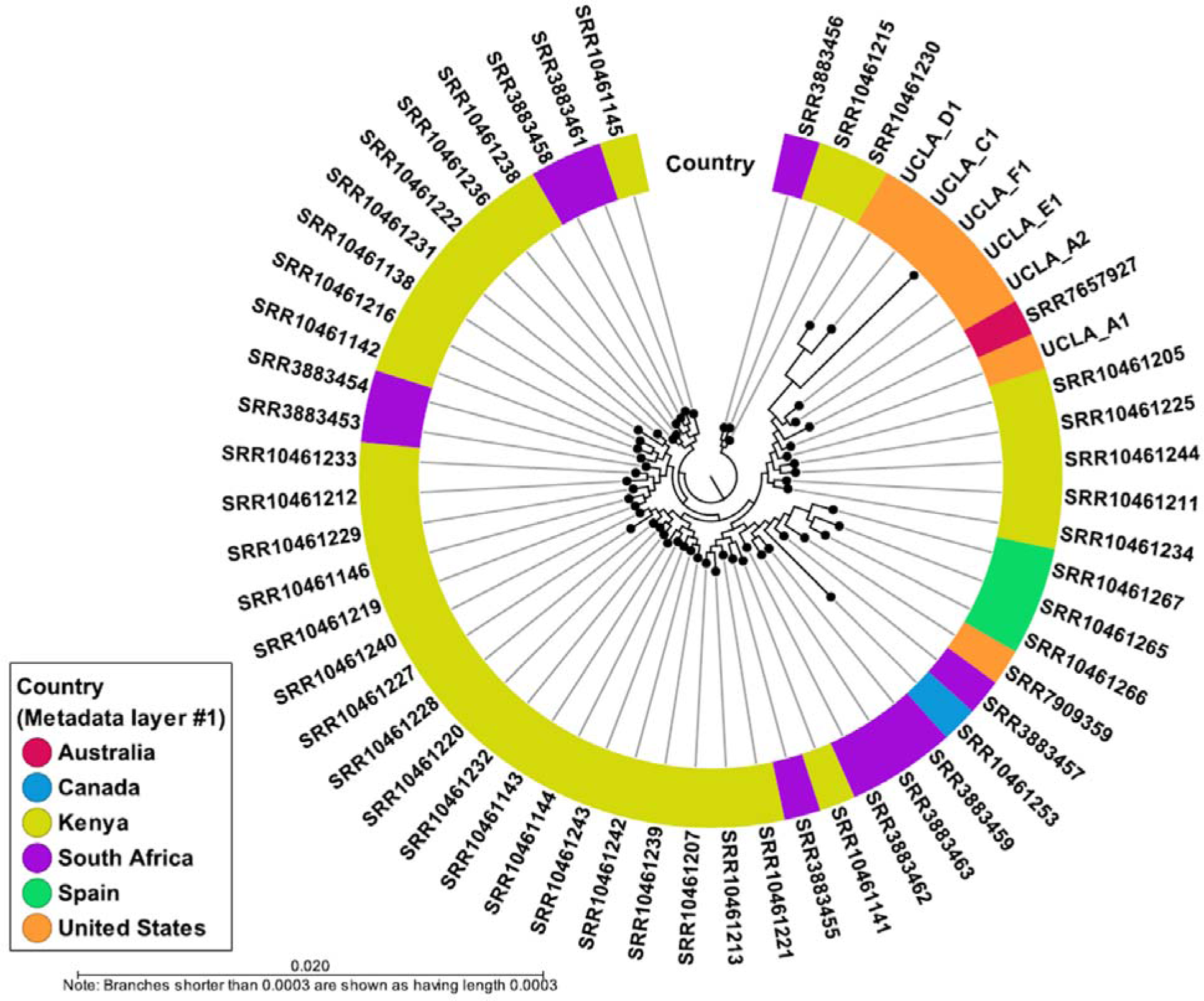
K-mer Analysis of 56 *C. auris* Isolates in Clade III. K-mer analysis was performed using CLC Genomics Workbench (Qiagen, Valencia, CA, USA) using genome sequences from 50 publicly available *C. auris* strains on NCBI GenBank as well as sequences from UCLA_A1, UCLA_A2, UCLA_C1, UCLA_D1, UCLA_E1, and UCLA_F1. A list of sequences used is found in Supplemental Data 1. The metadata (*i*.*e*., metadata layer #1) shows the country of origin.

